# Mothers’ experiences of using Facebook groups for local breastfeeding support: results of an online survey exploring midwife moderation

**DOI:** 10.1101/2022.06.04.22275948

**Authors:** H. Morse, A. Brown

## Abstract

**Problem:** The use of Breastfeeding Support Facebook (BSF) groups that are aimed at supporting women in specific local areas, with links to face to face support, is under researched. The role that midwives play in offering support to local mothers through these groups has not been examined.

**Background:** Access to ongoing support from professionals assists mothers in meeting their breastfeeding goals. Social media is increasingly being used to offer this support, improving maternal knowledge, self-efficacy and breastfeeding duration.

**Aim:** To examine impacts of moderator type on mothers’ perceptions of online breastfeeding support, including when midwives play an active role in moderation.

**Methods:** An online mixed methods survey was conducted in January 2020. Quantitative data was analysed descriptively and for associations using SPSSv26. Qualitative data was analysed thematically.

**Findings:** Two thousand and twenty-eight mothers who used local BSF groups completed the survey. The experiences of those participating in groups moderated by midwives versus other moderators such as peer supporters, were compared. Moderation was an important factor in mothers’ experiences, with trained support associated with greater engagement and more frequent visits, impacting on perceptions of group ethos, reliability and inclusivity. Midwife moderation was uncommon but valued, and associated with viewing local face to face midwifery support for breastfeeding more positively.

**Conclusion:** Midwife moderated or supported Facebook groups have the potential to add value to local face to face services and improve breastfeeding experiences in communities. The findings have important implications to support the development of integrated online interventions to improve public health.

**Statement of Significance:** *Problem:* Low breastfeeding rates are connected to inconsistent access to quality local support. Mothers are frequently turning to social media groups for support but little is known about who runs and moderates them.

*What is Already Known:* Mothers have concerns about trustworthiness and reliability, despite finding Facebook groups useful for shared experience. There is insufficient evidence on moderation to recommend online models within maternity services.

*What this Paper Adds:* Evidence that Facebook groups linked to local face to face support are valued, and that trained moderators improves experiences. Access to local midwife support within Facebook groups improves perceptions of local midwifery support overall.

## Introduction

Despite breastfeeding long being recognised as a public health priority, women encounter a variety of barriers to accessing the necessary ongoing professional support to meet their goals [1]. Although research has demonstrated the impact of consistent, high quality professional and social support upon successful breastfeeding [2]. increasing budget cuts to face to face support has left many women without the support they need and many maternity health professionals unable to give the level of face to face care they would like [3]. The recent COVID-19 pandemic exacerbated this issue [4].

Increasingly, to fill this gap, mothers are turning to social media to access the breastfeeding support they need [5,6] Facebook support groups have proliferated in recent years, and the platform is now commonly used by new mothers to access support during the transition to parenthood [7] including for specific aspects such as breastfeeding. Research has highlighted how mothers value this form of support [8] crediting it with increased breastfeeding duration [9].

Broadly there are two different types of breastfeeding support groups on Facebook. One type is a broad support group often with members from around the UK and internationally. Another more recent type is a local online Breastfeeding Support Facebook group (BSF) which is specifically linked to a local area (i.e. ‘Bridgend breastfeeding group’). Membership of the group often leads to contact with available face to face services. Recent research has highlighted that mothers find these groups useful, supportive and accessible, attributing them to helping them to breastfeed for longer [10].

It is clear that mothers who use Facebook for breastfeeding support value it. However it is not without its challenges. One core issue that often arises in social media breastfeeding support groups is the topic of group moderation. Moderators in social media groups monitor and regulate group posts to facilitate co-operation and prevent abuse [11]. They are essential to the success of online communities, maintaining activity and shaping the flow of discussion [12]. A lack of moderation of breastfeeding groups on Facebook has been highlighted as something mothers feel anxious about, either due to a lack of reliability of content posted or conflict arising between group members [13].

Many Facebook breastfeeding groups are based on a peer support format where other mothers who have breastfed, and may have received formal peer supporter training, lead and moderate discussions. Peer support is an effective method of helping mothers to continue to breastfeed [14]. offering solidarity and shared experience which are key to increasing breastfeeding knowledge and confidence [15]. However, access to qualified expertise is also central to high quality breastfeeding support, building trust and reassurance with evidence-based information [2]. Although many Facebook groups do have trained supporters or lactation consultant members and moderators, verifying and validating their expertise can present challenges. Mothers report concerns that Facebook groups are filling increasing gaps in NHS services, despite being unregulated [13]. They seek and value moderation they can trust to address misinformation and provide reassurance [8].

A second type of moderation of breastfeeding support on Facebook is through health professionals such as midwives. The rise of local BSF is likely one factor in encouraging this as it potentially enables midwives to reach local mothers through social media. However there is very little evidence examining the impact of this. Research examining other areas where midwives take on the moderator role in a Facebook group such as in pregnancy highlights how although there can be challenges [16] midwives valued being able to use their professional knowledge and communication skills to offer support, address misinformation and facilitate participation [17, 18]. This enables delivery of informational and emotional support simultaneously, breaking down traditional hierarchical barriers and encouraging service users to actively participate in their care [19]. The use of midwife moderators in pilot schemes has demonstrated success in providing supporter continuity and validated information [20; 18].

These benefits may apply to midwife moderation of local BSF groups, linking online and face to face breastfeeding support to widen access, offer reassurance and promote trust. The aim of the current study was therefore to explore mothers’ experiences in relation to lay, trained or professional moderation of local BSF groups, identifying any benefits and concerns. Understanding any value of midwife input to integrated online breastfeeding support will help support new approaches to using social media in practice, in education and to improve services.

## Methods

### Design

This study is part of a larger study exploring mothers experiences of using online local Facebook groups for breastfeeding support. This paper reports the findings specifically in relation to type of group moderation. The gathering of both quantitative and qualitative data via a mixed methods approach supported in depth description of both statistical relationships between BSF group provision, use and support needs and participants perspectives.

### Participants

Participants were mothers aged 18+, breastfeeding at least one baby up to 24 months old, and were currently a member of a local BSF group, of any moderator type. This was defined as a Facebook group identified as offering breastfeeding support to mothers residing within any specific geographic area within the UK, rather than to national or international members. UK postcodes were provided to confirm residency and analysed for distribution frequency. Exclusion criteria included age <18 years, inability to consent and inability to complete the questionnaire in English. Ethics approval was granted by a University Research Ethics Committee.

### Questionnaire design

A exploratory online survey, consisting of open and closed questions, was designed to enable large scale, efficient data collection. Questions were devised from the literature on midwife moderation, peer and online support [8, 13, 20]

Questionnaire items included:

- Demographic background e.g age, education, ethnicity
- Leadership and use of the local BSF group, e.g. who runs the group, how often mothers visit (parents, midwives, trained peer supporters, lactation specialists, unsure)
- Perceptions of breastfeeding support from midwives, e.g. face to face and online
- Awareness and experiences of online midwifery support
- Experiences relating to moderation e.g. lay, trained and professional

The questionnaire was piloted in a named local BSF group prior to sharing, and was completed by twelve mothers. Feedback from initial participants was positive on structure and content. No changes were required.

### Procedure

Data were collected using the online survey tool Qualtrics in January 2020, prior to the outbreak of COVID-19. Participants were recruited to the study via social media posts on Facebook and Twitter containing brief details of the study. Study adverts were also shared (after permission was sought from moderators) on local BSF groups across the UK. Adverts contained study information and inclusion criteria with a link to the participant information sheet for the questionnaire. If participants consented, the remainder of the questionnaire loaded. A short debrief was included at the end of the questionnaire with details of how to contact the research team or seek further support.

### Data Analysis

Quantitative questionnaire data was analysed using SPSS v26. Descriptive data was analysed for frequencies. Cross-tabulations, MANOVA and t-tests were used to explore associations and differences within the data, particularly how moderator type affected attitudes and experiences. Where indicated responses were compared using three moderator groups: unsure, midwife led and not midwife led (peer supporters, parents and lactation specialists). Thematic analysis was conducted to explore patterns and connections within the qualitative data. After familiarisation with the data, initial codes were produced, identifying themes which were reviewed in relation to the coded extracts, defined and named. These were reviewed by a second researcher and discussed until agreement reached (Braun & Clarke, 2006).

A reflexive journal was used to reflect on methodological decisions and the researcher’s background in breastfeeding support and influences as a health professional. Care was taken to avoid leading questions when designing the online surveys, instead offering a range of response options. Results were audited by the second researcher, providing feedback on the adequacy of data, development of findings and the interpretive perspective (Lincoln & Guba, 1985).

## Results

Two thousand and twenty-eight mothers completed the questionnaire. Mean age of participants was 32.35 (SD: 4.551; range 19 – 47). Mean age of infants was 10.6 months (SD: 6.393; range 1-24) (Table 1). Participants were members from 227 local BSF groups across the UK.

**Table 1:** Sample distribution by demographic factors.

| Indicator | Group | N | % |
| --- | --- | --- | --- |
| Age | ≤ 20 | 3 | 0.1 |
|  | 21-25 | 9 | 0.4 |
|  | 26-30 | 127 | 6.3 |
|  | 31-35 | 547 | 27.0 |
|  | 36-40 | 828 | 40.8 |
|  | 40 ≥ | 420 | 20.7 |
| Education | No formal | 11 | 0.6 |
|  | GCSE | 117 | 5.8 |
|  | A-Level | 307 | 15.2 |
|  | Degree | 883 | 43.9 |
|  | Postgraduate | 692 | 34.4 |
| Ethnicity | Asian/Asian British (Indian, Bangladeshi, Pakistani, Other) | 42 | 2.09 |
|  | Chinese | 5 | 0.25 |
|  | Black or Black British | 5 | 0.25 |
|  | Gypsy/Traveller | 1 | 0.05 |
|  | Irish | 35 | 1.74 |
|  | Mixed or multiple | 41 | 2.04 |
|  | Other | 12 | 0.60 |
|  | White/White British | 1872 | 93.0 |
| Marital Status | Married/civil partnership | 1451 | 72.2 |
|  | Divorced | 10 | 0.50 |
|  | Cohabiting | 474 | 23.6 |
|  | Single | 73 | 3.6 |
|  | Widowed | 2 | 0.10 |
| Employment | Full time | 819 | 40.8 |
|  | Part time | 828 | 41.2 |
|  | Not working | 361 | 18.0 |

### Leadership and moderation of UK local BSF groups

Participants were asked to indicate, via tick box of options (e.g. midwife, peer supporter, parents, lactation specialist), who provided group moderation of their local BSF group, with the option to reply ‘unsure’. Moderation was defined as taking responsibility for regulating posts and discussions. Trained peer supporters (47.9%), lactation consultants (29.1%) and parents (19.9%) made up the largest number of moderators. Overall, 5% of mothers reported belonging to a midwife moderated group. Some groups had mixed moderation across those categories. However, 20.7% of mothers were unaware of who provided moderation for the group. For the purposes of further analysis of midwife moderation, mothers were split into three main groups: midwife led (5%, n = 101), not midwife led (all other support), 75.4%, n = 1530) or unsure (19.6%, n=397).

Participants were asked how often each type of BSF group member/moderator (midwives, peer supporters, lactation specialists or other parents) provided them with support, and how useful they found it (Table 2). Midwife moderators offered a high level of support to mothers in their groups, with 87.5% having received midwife support often or sometimes and 97.8% rating this useful or very useful. Most mothers who did not have midwife input felt this would or may improve their experience of using the BSF group (84.3%).

**Table 2:** Participants who ‘often’/’very often’ received support from difference sources across different moderated groups.

| Group type | Midwife moderated |  | Not midwife moderated |  | Unsure who moderates |  |
| --- | --- | --- | --- | --- | --- | --- |
|  | N | % | N | % | N | % |
| Other parents | 84 | 94.4 | 1020 | 91.6 | 291 | 81.7 |
| Midwives | 77 | 87.5 | 475 | 42.6 | 157 | 44.1 |
| Peer supporters | 62 | 69.6 | 908 | 81.5 | 222 | 62.4 |
| Lactation specialists | 50 | 55.6 | 685 | 61.5 | 155 | 43.5 |

To understand whether moderation of the group had any association with frequency of using the group, participants were asked how often they used or visited the online group. Options were several times a day, at least once a day, a few times a week, one a fortnight or rarely. In a MANOVA, frequency of Facebook group use was associated with group moderator type [F (2, 1564) = 5.376, p = .005], with post hoc Bonferroni tests findings a significant difference between the midwife led and unsure groups (p=.0.006]. Mothers who belonged to group run by trained supporters visited most often.

#### Links between face to face and online BSF support

Participants were also asked whether they knew of any links between the BSF group and local face to face breastfeeding support groups. Overall, 67% mothers knew of a physical group linked to their online group and 69.8% of those had attended it. In terms of who led the face to face groups, lactation specialists (IBCLC) (23.3%) and trained peer supporters (37.6%) provided most face to face support groups. Conversely, few face to face groups were led by midwives (4.8%). Some groups had mixed leadership across those categories.

Overall, 541 (26.7%) had also met a trained (midwife, peer support, lactation specialist) BSF group moderator in person, demonstrating evidence of continuity of care/support. This was particularly true for those using midwife moderated groups, where 48% had met those running the BSF group, including 14% who had also received clinical care from them. See Table 3 for details.

**Table 3:** Frequency of continuity of in person and online support by group type.

| Group type | Midwife moderated |  | Peer supporter moderated |  | Lactation specialist moderated |  |
| --- | --- | --- | --- | --- | --- | --- |
|  | N | % | N | % | N | % |
| Met at a face-to-face support group | 34 | 34 | 313 | 32.2 | 187 | 31.6 |
| Received clinical/ one to one care | 14 | 14 | 43 | 4.4 | 24 | 4.1 |
| **Total** | **48** | **48** | **356** | **36.6** | **211** | **35.7** |

The association between perceptions of online and face to face support was also explored. First, participants were asked how well supported they felt they had been by a range of offline local sources, including friends, family and health professionals. Overall, 1539 (88%) of mothers felt they had been well supported by midwives face to face. Perceptions of support were compared for those in a group moderated by midwives versus those in a group not moderated by midwives/unsure. Significantly more mothers who had access to midwife support on Facebook felt well supported by local midwives than those who did not have online midwife moderated support [t (1746) = -3.105, p = .003]. Overall, 82 (81.2%) mothers in midwife moderated groups agreed that the personal, offline support for breastfeeding they received from midwives was positive, compared to 1083 (69.7%) of those in the lactation consultant/peer support moderated groups (Table 4).

**Table 4:** Perceptions of positive local midwifery support for breastfeeding by group type (showing proportion of participants who strongly agree / agree)

| **Source** | **N** | **%** |
| --- | --- | --- |
| Midwife led | 82 | 81.2 |
| Health Visitor led | 10 | 62.5 |
| Parent led | 282 | 70.3 |
| Lactation specialist/peer support | 1083 | 69.7 |

### Impact of moderator type on mothers’ experiences

Mothers rated their agreement (strongly agree to strongly disagree via a 5 point Likert scale) with a series of questions about their perceptions of their online BSF group. Overall, 14.5% felt it gave them additional access to midwifery support unavailable elsewhere, and most mothers agreed that their BSF group was a reliable source of information overall (91.2%). However, when the three groups, midwife led, non-midwife led and unsure were compared, moderation type affected perceptions (Table 5).

**Table 5:** Moderator related BSF group experiences by group type.

|  | Midwife | Not midwife | Unsure |
| --- | --- | --- | --- |

|  | moderated |  | moderated |  |  |  |
| --- | --- | --- | --- | --- | --- | --- |
|  | N | % | N | % | N | % |
| Confidence in reliability of group information | 85 | 93.4 | 1033 | 92.6 | 311 | 86.4 |
| Midwife contributions improve confidence in the advice | 67 | 73.6 | 296 | 26.5 | 150 | 41.7 |
| Reassured by access to trained support on group | 86 | 94.5 | 986 | 88.4 | 241 | 66.9 |

A MANOVA was used to explore differences in experiences between the three types of Facebook group (midwife led, not midwife led or unsure). A significant difference was found between the three types of groups for:

‘I feel confident that the information on the group is reliable’ [F (2, 1549) = 15.713, p = .000]. Post hoc bonferroni tests showed a significant difference between the midwife and unsure group (p = .000) and not midwife led and unsure group (p = .000) but not between the midwife led or not midwife led groups (p = .705). Both the midwife led and not midwife led groups reported more confidence than the unsure group.

‘I feel more confident taking advice if midwives add to the discussion’ [F (2, 1549) = 63.339, p = .000]. Post hoc bonferroni tests showed a significant difference between all groups (p =.000). Those in the midwife group felt the most confident, followed by those in the unsure group and finally those in the not midwife led group. Large differences were seen in mean scores with those in the midwife group on average agreeing or strongly agreeing whilst those in the not midwife group averaging between neutral or disagreeing.

### Exploring experiences of moderation

Participants were asked to use open ended boxes to further reflect on their experiences of accessing local breastfeeding support online. Moderator type and approach, and the siting of the BSF group within local services, was key to many positive and negative experiences. These included perceptions of reliability, trust and inclusivity that mothers associated with the value of the support, and a sense of belonging to the on and offline community. Thematic analysis of experiences identified four themes: Local group format, Moderation, Confidence in expertise and Trust.

#### 1. Local group format

Mothers highly valued the local group format, describing a variety of advantages and ways in which it had enabled them to access support. Two sub themes arose, describing the role of the group and its links to local services in supporting mothers to build relationships with, and confidence in, group members and moderators.

##### a) Overcoming barriers

Mothers described how the online group being situated within, or connected to, local services helped them to overcome barriers to making connections and seeking support. For some mothers the BSF group provided reassurance that local help was accessible and available if needed.

> *‘I suffered with terrible postnatal depression and never felt I could attend in person but I liked knowing I had support close by*.*’* (Aged 25-30, baby 0-6 months)

Overcoming barriers to seeking face to face support was particularly important for those with younger babies, often feeling overwhelmed and in need of support but not yet ready or able to attend a group. Online support was seen as offering access to support, practical or emotional, in a convenient and self-directed format and ‘building a bridge’ to developing relationships with local parents prior to physically meeting them.

> *‘Sometimes people underestimate the impact of being home alone in those early days, whilst also not being physically/emotionally able to get out, and possibly not wanted to, but still needing support. Also with issues with anxiety the idea of going to face to face group whether it is social or NHS service etc can feel overwhelming, like you will be judged or you will have to be friendly or have more energy than you do etc whereas the online support takes these potential issues away then helps you feel connected. Whilst giving the opportunity to talk to like-minded people and be able to attend a face to face group if/when you feel ready and may know some of them by then*.*’* (Age 30-35, baby 6-12 months)

##### b) Linking services

Mothers valued access to trained support that was linked across the online group and face to face groups, enabling them to validate information and build trusting relationships with members and moderators. This interaction built confidence and facilitated ‘real life’ social connections, enhancing the concept of ‘building bridges’.

> ‘*As it’s local, you sometimes recognise other mums from the online group when you then meet them in person at a breastfeeding group / other baby group. Really supportive environment which has made me feel much more confident and able. You are often braver asking things online than face to face. Again, as it’s local there are often invites for play dates / meet ups so you can get to know other mums if you are feeling isolated*.*’*
>
> (Aged 30-35, baby 12-24 months)

Mothers particularly valued local health professionals working alongside others to provide this, supporting their confidence in the expertise available locally and generating positive regard for the service as a whole.

> ‘*Well moderated group by health visitors and trained peer supporters. Great link to physical group, often people come to group after seeing positive support on fb group. Great way to share up to date articles/videos, peer supporters often post resources and links following discussions at groups. It’s a really positive fb group and everyone’s really supportive when you post questions, peer supporters or HV team respond to most posts and will encourage face to face meets when needed e*.*g. latching issues*.*’* (Aged 30-35, baby 12-18 months)

#### 2. Moderation

Mothers felt that their local BSF group was a virtual community that they valued for access to information, emotional support and solidarity, and that this was well facilitated by all types of moderator. However, some described examples of moderation within local BSF groups that they felt was negative, defensive or unpredictable, undermining trust in the online community and the local support available on and offline.

##### a) Group ethos

Some mothers described negative experiences of moderation, often associated with parenting styles, that had impacted the sense of inclusivity and belonging that they valued. Others felt some groups actively damaged relationships between mothers and local health professionals, highlighting the importance of improved links and opportunities for meaningful online interaction with care providers.

> *‘Our local breastfeeding support group is moderated by very biased individuals. They have set a very strict group ethos and will only let you discuss things that suit their parenting style. They often do not use evidence-based advice. There is an awful lot of negative behaviour towards health care professionals from both parents and moderators which increases the risk of mothers not accessing help when/ if needed. The moderators do not actively try to improve the relationship between mothers and HCPs*.*’* (Aged 36-40, baby 6-12 months)

Midwife moderation was described by one mother as a solution to overcoming issues that prevented her from feeling able to contribute. Group ethos was a powerful positive or negative driver of belonging, linked again to improved connections with providers.

> *‘I wish I could become more actively involved in the local support group but unfortunately my lifestyle choices and parenting choices do not match the moderators so I don’t feel I can contribute. I would love for the group to be regulated by midwives or other HCPs*.*’* (Aged 31-35, baby 6-12 months)

##### b) Health professional input

Mothers highly valued the input from health professionals where available, describing positive impacts from access to rapid, validated support and an appreciation for the service as a local resource. Online support from midwives or health visitors was associated with higher regard for local maternity services as a whole.

> ‘*Huge amount of support from highly trained, knowledgeable professionals. Much more helpful than information and help received from other services…Extremely fast responses. And people are clearly passionate about breastfeeding and supporting mothers to do so. One of the best resources available*.*’* (Aged 25-30, baby 6-12 months)

However, there were also negative experiences of health professional input, particularly where the support group was not provided by them but used defensively, rather than to offer support. Mothers’ identified the need for open and honest interactions, linked with group ethos and moderation type/style.

> ‘*Only time I have seen a professional use the group it has been only to comment on a post that has been complaining about midwives in order to defend another midwife’s actions or explain a local NHS policy for example*.’ (Aged 25-30, baby 12-18 months)

#### 3. Confidence in expertise

The notion of health professional input was valued by all mothers and overall, mothers not already using midwife moderated groups felt this added provision would be of benefit. However, some mothers had concerns about expertise, poor experiences of midwife support and a desire for collaboration to ensure adequate skills and accessibility.

##### a) Experiences of offline support

Some mothers described concerns about the level of breastfeeding training received by midwives, including examples of a lack of knowledge and receiving inaccurate or out of date information. Others felt breastfeeding knowledge came from personal experience professionals may not have.

> *‘I have found Health visitors and midwives advice very misleading and poor. I recommend the group because I want people to get advice from experience rather than what people “learn” from a book’* (Aged 25-30, baby 12-18 months)

Some mothers felt their confidence in midwives’ ability to provide high quality breastfeeding support had been undermined by poor experiences. These mothers questioned whether midwives would be able to provide them with quality online support.

> *‘Midwives input would only be helpful if they have extra training on supporting breastfeeding. My experience was that some had patchy or outdated knowledge*.’ (Aged 30-35, baby 12-18 months)

##### b) Valuing moderator expertise

However, it was more common for mothers to describe a lack of professional or trained support as a downside to a group, seeking expert input to validate information and offer reassurance.

> *‘[A negative of the group is] no peer supporter or midwife. Only parents’ experiences and sometimes advice can be outdated and go against NHS and WHO guidelines*.*’* (Aged 30-35, baby 6-12 months)

Even where mothers described professional face to face support as lacking, expert input was seen as fundamental to online support. This was particularly valued if BSF group input and moderation demonstrated collaboration between local health professionals and peer support.

> *‘My experience of midwife, HV and GP was that they could be hit and miss in terms of availability and knowledge. Having a community of mothers with direct experience, moderated by [lactation] experts very helpful*…*’* (Aged 30-35, baby 19-24 months)

#### 4. Trust

Online groups are simple and free to set up and mothers raised issues of how to assess the credentials of those providing them and concerns about validating the information offered. Two sub themes arose from mothers’ awareness of who was offering support and their perceptions of this affecting its reliability, developing or undermining trust in their group.

##### a) Verifying support

Some mothers described being unable to easily verify the source of support or group moderator. Identifying credentials or training was of concern in groups not associated with local health services.

> *‘No guarantee that someone who says they are a professional (e*.*g. a midwife or breastfeeding counsellor) actually IS a professional*.*’* (Aged 25-30, baby 12-18 months)

##### b) Regulation and moderation concerns

A lack of professional or trained moderation was associated with concerns about the reliability of advice and the ability to appropriately signpost to face to face services.

> *‘Unqualified advice where many posts are not answered by a trained peer support worker and the responses are not governed by them either. I have never seen members of the group express a concern to a potential breastfeeding problem or suggest the original post author seeks qualified support. Every issue receives the blanket “its normal” response*.*’* (Aged 30-35, baby 19-24 months)

Some mothers also associated a lack of regulation of BSF groups with wider issues of confidentiality, communication skills and safeguarding. This sub theme strengthened the points made in relation to confidence being built through linked services and verified expertise.

> *‘Reliability of who is actually part of the group, confidentiality of issues raised and privacy if you know people in the group in the real world*.*’* (Aged 30-35, baby 12-18 months)
>
> *‘I am a trained peer supporter and feel some ‘advice’ which although well-meaning can be wrong, less than tactful and in some cases could be dangerous*.’ (Aged 25-30, baby 0-6 months)

## Discussion

This study explored women’s experiences of belonging to local BSF groups, specifically how they are moderated, by whom and what the role of midwives is in this. There is a growing body of research that shows the value of online breastfeeding support [8,15] including local groups [10] but little evidence of the impact of professional moderation that may be linked to maternity services and other face to face local support. Our findings explore differences in moderation and moderator type to identify any impact on mothers’ experience of online support. The findings have important implications for the potential delivery of localised breastfeeding support by health professionals and within maternity services.

Local BSF groups are now widely used by breastfeeding mothers across the UK and have provided vital access to support during the suspension of many face to face services during the COVID-19 pandemic [4]. Although support strategies that are weighted towards face-to-face support are more likely to successfully increase rates of continued exclusive breastfeeding [2], online support can offer a range of additional benefits [21, 15]. Mothers credit the convenient, timely access to information and solidarity with supporting them to meet and exceed their breastfeeding goals [9]. Using social media, mothers can offer each other support around the clock, overcoming physical and emotional barriers to attending face to face support groups, within a trusted community [10].

However, face to face support remains important and repeated systematic reviews show successful breastfeeding support interventions offer provision across a combination of settings (hospital, home and community) [22, 2]. One of the core findings of our research is that local online BSF groups often provide a link or gateway between social media groups and face to face groups [10]. Feeling connected to the group, seeing familiar faces and trusting information appeared to strengthen the likelihood that some women would go on to attend face to face groups. Mothers valued the links between on and offline support for access to shared experience, local knowledge and the opportunity to build ‘real life’ supportive relationships with members and moderators. These factors are key to building a support community that, by existing both on and offline, can normalise breastfeeding to improve mothers’ experiences and extend breastfeeding duration.

Research into the delivery of online support has identified that the majority is provided by charities or volunteers [15]. Our findings echo this, most local BSF groups were provided by other parents, peer supporters and lactation consultants with little health professional input. Whilst mothers highly value these volunteer-run groups for their lived experience and information sharing, they also perceived them as filling a gap in the support that health professionals and NHS services are failing to provide [13; 15]. Social support is a critical element of both initiating and sustaining breastfeeding, and access to a local BSF group (and linked face to face group where available) provides mothers with access to the knowledge and experiences of other mothers [23]. However, accessing this online poses challenges for mothers in validating the information shared and the expertise of those providing it [13].

Moderation is a key feature of online communities [24]. Moderators or group administrators enforce guidelines and address misinformation, ensuring discussion remains respectful and constructive [11]. Unlike other health services, mothers are uniquely reliant on peer support for practical information to make breastfeeding work [25] so the accuracy of information and quality of support is essential [26]. Evidence suggests that without trained moderators, discussion in peer (mother led) communities for pregnancy can be inaccurate, lacking in credible evidence base and potentially harmful [27]. Addressing inaccuracy and offering effective support whilst maintaining a respectful community is a skill developed by moderators [24]. Mothers identify effective moderators as having as a proactive, facilitative and authentic approach, one which both offers accurate information and supports the development of trusting relationships [28]. This was echoed in our findings; poor experiences of moderation left women feeling judged, unwelcome or not able to ask for support. Professional-led or moderated local BSF groups may therefore provide a solution to varying moderation and validating expertise.

The use of midwife moderators for pregnancy and postnatal Facebook support has demonstrated success, meeting mothers’ support needs whilst providing professional access to midwives [29]. Offering support in private Facebook groups, moderated by midwives, the ‘Facemums’ initiative provided improved supporter continuity and access to validated information [17, 20]. Conversely, other studies have found peer-led BSF groups are preferred over professional-led groups, who described professional online support as impersonal and unsympathetic [30]. Our findings reflect that mothers found midwife moderation and subsequent support useful and valuable. Knowing support was available from midwives increased confidence in taking advice and perceptions of reliability, whilst mothers who were unsure who was responsible for moderating the group they belong to, who used it less and perceived it as less reliable.

Although in our study mothers who had access to midwife moderated BSF groups were in the minority (5%), we found they were highly valued by those who did have access and sought after by those who did not. However, some mothers did have concerns about whether all midwives were suited to this role. Concerns centred on experiences of receiving inaccurate or out of date information from midwives face to face, which had undermined their confidence in the ability and commitment of midwives to providing high quality breastfeeding support. Conflicting and inconsistent advice and approaches to breastfeeding support from midwives that mothers find pressurising or undermining are common experiences [31]. Midwives experience barriers to providing the individualised care women need, including time and staffing pressures, negative personal experiences and workplace culture [32]. Most midwives want to provide woman-centred, individualised care [33] and are unhappy with the standard of breastfeeding support they feel able to provide [34]. However, there are also concerns about the ability of standard midwifery training to challenge the negative attitudes and knowledge gaps that affect women’s care and breastfeeding experiences [35].

A key question is therefore how we can better support midwives to develop knowledge and skills around supporting breastfeeding, which is important not just for providing social media support but breastfeeding care more generally. To offer effective breastfeeding support, midwives need embodied knowledge, developed via vicarious and work based experiences, as well as cognitive knowledge [35]. Moderating Facebook groups offers midwives a ‘window into women’s experiences’ [29]. Involvement can facilitate reflexive and reflective practice development in a sustainable way [17], similar to the successful biopsychosocial approach used to train peer supporters, which results in high levels of knowledge and positive attitudes [36]. The BSF format therefore offers opportunities to improve practice as well as support provision.

To meet their breastfeeding goals, mothers need to feel supported and respected in their choices by maternity services, yet a 60% of UK mothers report receiving little or no support from midwives [37, 34] Around a fifth seek to fill this gap elsewhere, including seeking online support [34]. Notably we found there was an association between feeling supported by midwives online and offline. Mothers in BSF groups moderated by midwives were more likely to feel that midwives they saw face to face were supportive of breastfeeding. These improved perceptions suggest that online provision can enhance support for mothers and feedback for services, a clear benefit of integrated, local midwife moderated online support.

Involvement in online support communities also offers midwives greater immersion in women’s experiences and provides opportunities to engage actively with evidence. These experiences increase midwives’ knowledge and understanding with a direct and positive impact on their practice [29, 18]. So, moving forward, how do we enable more midwives to use their skills online in supporting mothers through this format?

We need to recognise the benefit to mothers, midwives and maternity services in engaging with social media support, addressing the mixed messages between national guidance and employer policies [38, 18] to promote safe and effective use. NMC social media guidance lists building relationships with service users as a risk to midwives’ professional registration, without differentiating between personal and professional use [39]. This distinction is clearly needed to support midwives to offer this support. We are doing further research to explore in depth the barriers and facilitators to greater midwife involvement, but it is clear that online breastfeeding support needs cannot be met by midwives alone.

Collaboration with peer supporters, lactation consultants and third sector providers in providing breastfeeding support is known to ensure the best outcomes for mothers [2, 37]. Whilst we found mothers highly valued midwife moderation when it was available, they also described shared experience as central to what they sought from an online BSF group. Mothers emphasised the value of peer support from mothers at varying stages of breastfeeding, enabling them to offer support to those with younger babies and gain insights from those with older ones. Providing support beyond early infancy may fall outside midwives’ education, experience and scope of practice.

Mothers recognised this, identifying the depth of knowledge offered by lactation consultants and lived experience of other mothers and peer supporters as important features. Trained peer supporters bring experiential knowledge to the community, alongside high levels of breastfeeding knowledge and, significantly, more positive breastfeeding attitudes than health professionals [36]. However, knowing the group was linked to local midwife support had clear benefits in establishing the group as a reliable, trusted source of support. Collaborative services are positively evaluated by midwives, mothers and peer supporters [40, 18] but require clear roles and responsibilities and shared working to promote integration and enhance cohesion [25, 18]. Further research needs to identify how this can be applied to local BSF group provision.

Burnout and stress, arising from increasing workloads and reduced resources, is a significant issue amongst the UK midwifery workforce [41]. Recommendations for participation and development of digital roles must consider the implications for scope of practice, and the appropriate allocation of training, resources and renumeration to prevent BSF support becoming additional, unpaid workload. However, there are also significant potential positive outcomes for midwife moderators [18, 20]. Engaging in an online community improves continuity and collegiality [17] known to improve midwives’ job satisfaction and wellbeing, resulting in better care for mothers [43]. A growing body of evidence suggests there may be additional benefits to midwives, including greater confidence in knowledge, opportunities for reflection and improved communication [18]. The ‘Facemums’ model of local midwife moderated private Facebook groups identified that this model of support can be implemented in a scalable and sustainable way [17, 20, 29]. Further study is needed to explore the barriers and facilitators to applying this to breastfeeding support.

## Conclusion

Our findings suggest an emerging evidence base for the development of local BSF group formats by maternity services, in collaboration with peer support services. However, further research is needed to explore the scope of the role and the training, support and investment needed.

The research does have limitations. Mothers were current members of BSF groups, so the views of those with negative or ambivalent experiences and reasons for leaving are not captured. Mothers were older, with a high education level than average, although this is reflective of those in the UK who breastfeed for longer [44]. Participants were self-selecting and more likely to represent the most motivated to take part. However, the study advertisement asked for participants using all local BSF groups, so participation was not skewed by moderator type, and many described positive perceptions of a range of face to face support. Data was also collected prior to the outbreak of COVID-19, which increased online support seeking, including from health professionals, and it is likely the results would be affected by this context.

Our sample was also predominantly from White or White British backgrounds (93%). This may be as a result of self-selecting methods often underrecruiting those from ethnic minority backgrounds, but may also reflect barriers to participation including a lack of diversity in BSF leadership [45]. Stark disparities in maternity care and outcomes [46] may impact perceptions of health professional support among women from ethnic minority backgrounds. Findings should therefore be treated with caution but do provide insight into the experiences of women using these groups for support, and their perceptions of professional input.

Limitations aside, the findings are important in highlighting the widespread use of BSF groups, the specific value of locally linked on and offline services and issues of moderation. Whilst midwife moderation is not common, it has positive impacts on mothers’ perceptions and experiences, worthy of consideration when commissioning services. Findings support the development of clear frameworks for midwife moderation, and further research into collaborative provision to improve services. Future study is planned to explore midwives’ perspectives, identifying potential impacts, barriers and facilitators to the development of local BSF group provision.

## Data Availability

All data produced in the present study are available upon reasonable request to the authors

